# Rapid SARS-CoV-2 variants spread detected in France using specific RT-PCR testing

**DOI:** 10.1101/2021.02.20.21251927

**Authors:** Stéphanie Haim-Boukobza, Benedicte Roquebert, Sabine Trombert-Paolantoni, Emmanuel Lecorche, Laura Verdurme, Vincent Foulongne, Christian Selinger, Yannis Michalakis, Mircea T. Sofonea, Samuel Alizon

**Author notes:** equal contribution (Mircea T. Sofonea, Samuel Alizon).

## Abstract

SARS-CoV-2 variants raise major concerns regarding the control of COVID-19 epidemics. We analyse 40,000 specific RT-PCR tests performed on SARS-CoV-2-positive samples collected between Jan 26 and Feb 16, 2021. We find a high transmission advantage of variants and show that their spread in the country is more advanced than anticipated.

## 1 Context

Since the end of 2020, at least three SARS-CoV-2 strains, also referred to as ‘variants’, bearing an unusually high number of mutations have been associated with rapid epidemic spread in the UK (lineage B.1.1.7 [1]), South-Africa (lineage B.1.153 [2]), and Brazil (lineage P.1 [3]). Because of their increased transmissibility [4, 5, 6] and potential ability to evade host immunity [7, 8], monitoring these variants is important in the context of mass-vaccination. In France, since Feb 5, 2020, every positive RT-PCR test is tested with an additional PCR with probes targeting the ∆69-70 deletion and the N501Y mutation, both in the Spike gene. Lineage B.1.1.7 is associated with the two targets being detected. For lineages B.1.153 and P.1, only the N501Y mutation is present. If only the deletion ∆69-70 is detected, the infection may be caused by another variant or by a wild type strain with a deletion. Finally, if both targets are not detected, the host is considered as being infected by a ‘wild type’ strain. These tests are cheaper and easier to implement than full genome sequencing, which allows for their rapid deployment at a wide scale.

## 2 Factors associated with variant detection

We report the results of RT-PCR testing for SARS-CoV-2 strains using two techniques, VirSNiP SARS-CoV-2 Spike del+501 (TIB MOLBIOL) and IDTM SARS-CoV-2/UK/SA Variant Triplex (ID SOLUTION), both of which contain two probes that target the ∆69-70 deletion and the N501Y mutation.

Tests were performed on 42,229 positive samples collected between Jan 26 and Feb 16, 2021, on 40,777 individuals from 12 French regions. Some of the samples (3,323, i.e. 7.9%) originate from hospitals (mostly hospitalised patients) but the vast majority originates from the general population. 1,397 patients had multiple testing and only their first test was kept in the analysis. We also only kept data from individuals aged from 5 to 80 years old to minimise the weight of pre-school children and aged care facilities in the analysis. We also removed individuals with unknown age or region of testing.

Overall, we analysed 35,208 SARS-CoV-2-positive samples from the same number of individuals. 6,702 (19%) variant-specific RT-PCRs were uninterpretable because of an insufficient amplification of the internal control (ID solution) or of the targets (Tib MOLBIOL). These results were treated as missing in the analyses. Given that most of the variant were B.1.1.7 (Figure S3), we grouped samples that were B.1.1.7, B.1.53, and P.1-positive into a broader class of variant-positive.

We used a generalised linear model (GLM) to analyse the binary strain variable (with values ‘wild-type’ or ‘variant’). The explanatory variables were the patient age, the RT-PCR kit for variant detection used, the sampling date, and the sampling region. Further details are provided in Supplementary Methods.

Using a type-II analysis-of-variance, we find that all factors except the RT-PCR kit are significantly associated with the presence of variants (Table 1). In particular, the proportion of variants increased with date and decreased with age (Figure S1) and with hospital samples.

**Table 1:** Odds ratios and 95% confidence interval (CI) for the general model.

|  | Odds Ratio | 2.5% CI | 97.5% CI |
| --- | --- | --- | --- |
| date (per day) | **1.07** | 1.03 | 1.11 |
| age (per year) | **0.993** | 0.992 | 0.995 |
| kit:2 | 0.94 | 0.93 | 1.16 |
| location:non-hospital | **1.25** | 1.13 | 1.39 |

To investigate the temporal trends, we fitted a logistic growth model to the fitted values of an analogous GLM only on the data sampled outside hospitals (see Supplementary Methods). Assuming that variations in frequencies are driven by transmission advantages, we find that variants have a 50% (95% CI [37,64]) transmission advantage over ‘wild type’ strains (Figure 1). If all uninterpretable tests are assumed to be ‘wild type’ instead of being neglected, this advantage is 36% (95% CI [26,48]) (Figure S2).

**Figure 1:**
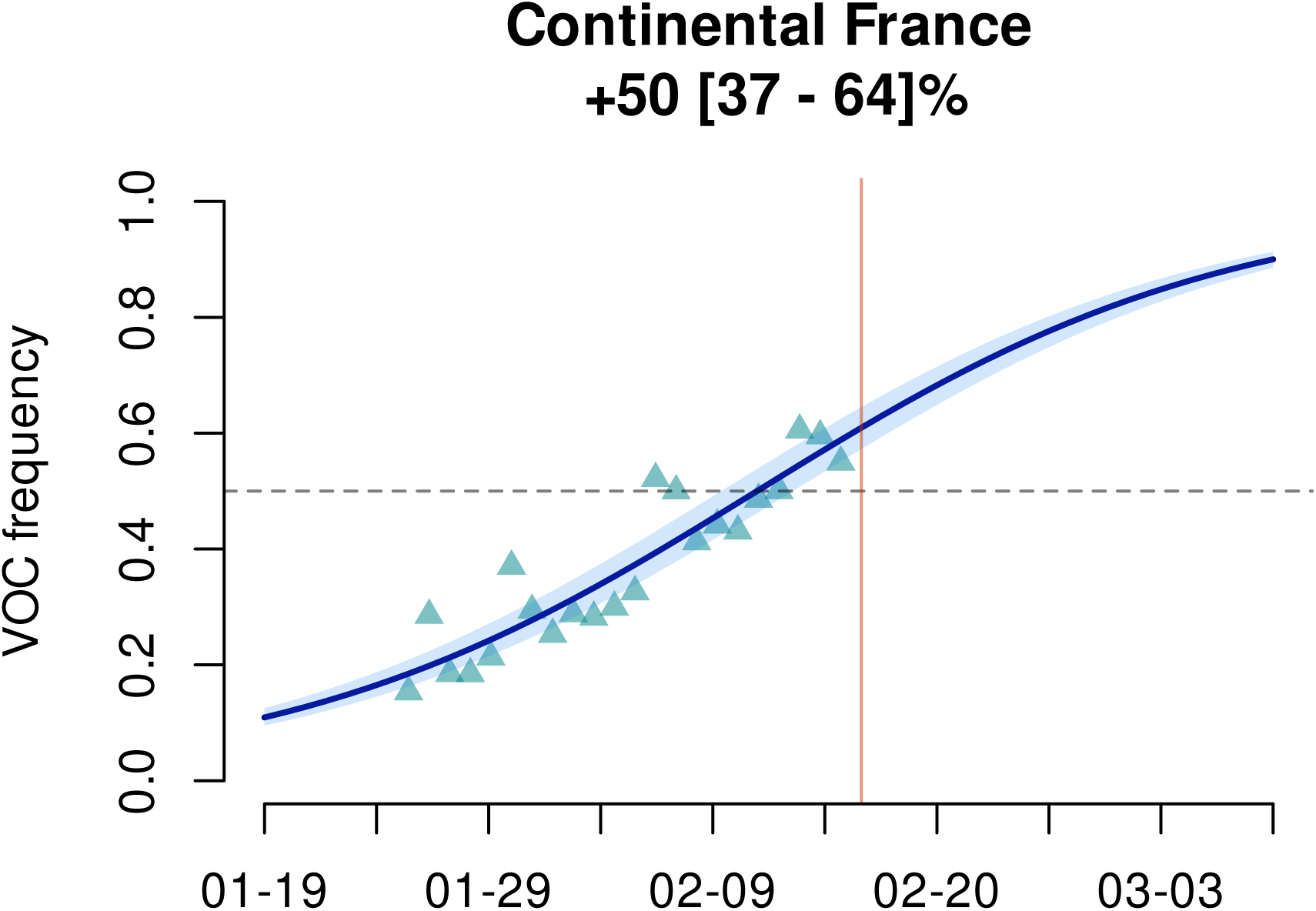
Estimated variants frequency kinetics in France. The triangles indicate the GLM-fitted values and the line is the output of the logistic growth model estimation. The vertical orange bar indicates the date of the analysis. The top figures indicate the estimated transmission advantage (ETA) of the variants (with respect to the wild type reproduction number) and its 95%-confidence interval (see the Supplementary Methods more details).

## 3 Regional analyses

The analysis-of-variance already showed that variant frequency varied across regions (Table 1).

We performed the logistic growth fit at the local level for the regions where enough data was available. Results are shown in Figure 2. We find that the growth advantage of the variant is more pronounced in some regions. We also find that in some regions, such as Ile-de-France, nearly half of the infections already appear to be caused by the variants, whereas in others, such as Burgundy, this proportion will not be reached until March 2021. However, some regions are less sampled in this analysis, which could affect local estimates.

**Figure 2:**
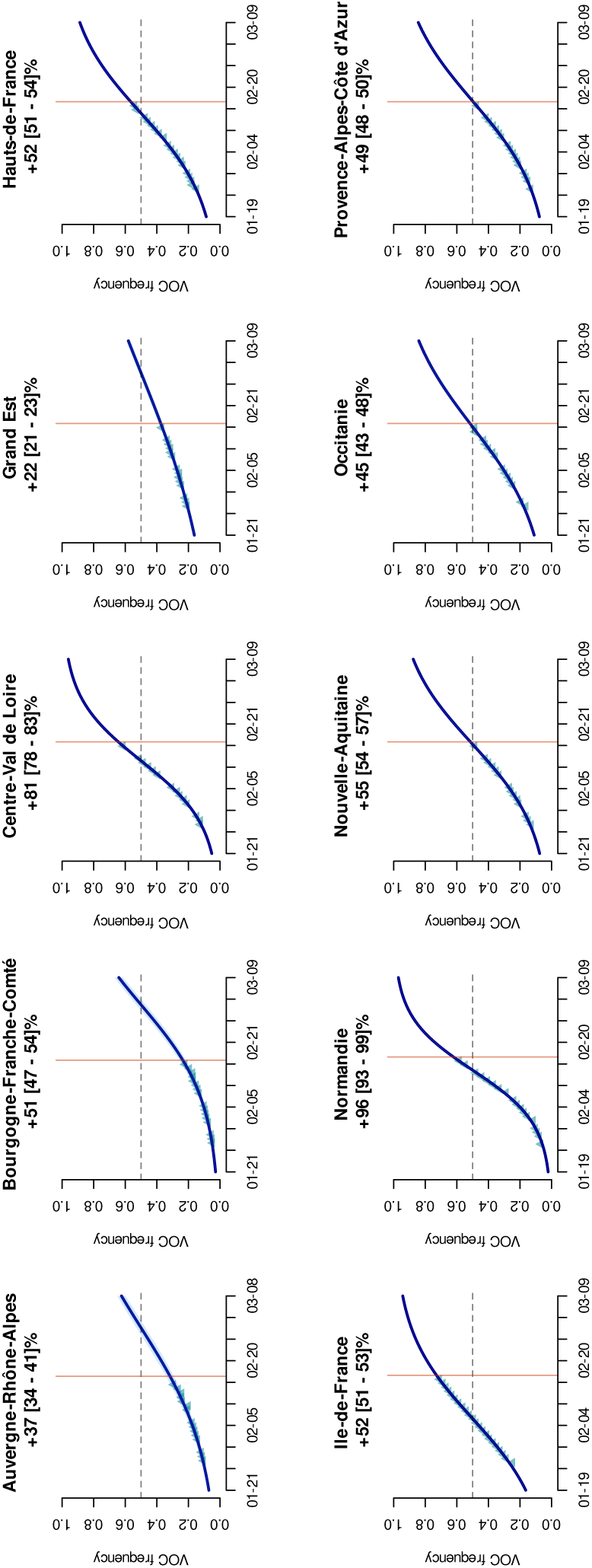
Logistic growth at the regional level. The dots indicate the GLM-fitted values values and the line is the output of the logisitic growth model estimation. The vertical orange bar indicates the date of the analysis. The top figures indicate the estimated transmission advantage (ETA) of the variants (with respect to the wild type reproduction number) and its 95%-confidence interval (see the Supplementary Methods for additional details).

## 4 Epidemic reproduction number

Finally, we investigated the correlation between the increase in the frequency of variant detection amongst positive tests in a region and the temporal reproduction number, denoted ℛ_*t*_, in that same region. The latter was estimated from COVID-19 ICU-admission data using the EpiEstim method [9] with a serial interval from [10], as described in [11]. Further details are provided in the Supplementary Methods.

We find a positive trend but this is not significant (Spearman’s rank correlation test, *ρ* = 0.30, p-value=0.34, Figure S4).

## 5 Discussion

We used two variant-specific RT-PCR tests to detect the fraction of infections caused by SARS-CoV-2 B.1.1.7, B.153, and P.1 in French regions between Jan 25 and Feb 16 2021. Our results have several practical implications. That we did not find any significant difference between the two specific RT-PCR kits used suggesting that similar data collected in France could be pooled.

In general, we find that many infections screened are caused by variants, especially B.1.1.7, with an increasing trend over time. Based on our estimates, more than half of SARS-CoV-2 infections in France could already be caused by variants, although there is some level of spatial heterogeneity. In a conservative scenario, where all uninterpretable tests are assumed to be caused by the wild type instead of being treated as missing values, a majority of infections should be caused by variants by the end of week 7 of 2021.

Variant-positive samples originate from significantly younger patients. This could be generated by some underlying stratification non-accounted for in this data. This is also consistent with earlier reports [4, 5], which also found the increase in variant proportion to be associated with higher basic reproduction number [4, 5]. We do find such a trend among French regions but it is not statistically significant.

A limitation of this study is that in spite of its density, sampling was performed retrospectively. This could generate biases if, for instance, transmission chains associated with variants are increasingly sampled. However, we do know if the sample was performed in a hospital and find that these are associated with a lower probability to detect a variant. This is consistent with the 14-day delay between infection and hospitalisation [12]. Another limitation is that specific RT-PCR does not have the resolution of full genome sequencing and other variants of concern may be underestimated or missed with this approach. However, the time scale considered and the relatively slow evolutionary rate of SARS-CoV-2 make this approach appropriate to monitor variant spread. Furthermore, it allows for exhaustive and timely testing of all the positive tests.

These results illustrate that variant-specific RT-PCRs are an interesting option for SARS-CoV-2 epidemic monitoring because of their cheap price and rapid deployment. They also reveal that the spread of B.1.1.7, B.1.153, and P.1 SARS-CoV-2 variants in France is faster than predicted [13], although with strong spatial heterogeneity, which stresses the importance of swift public health responses through vaccination and non-pharmaceutical interventions.

## Data Availability

Data will be made available upon manuscript publication

## Acknowledgments

We thank the ETE modelling team and Florence Débarre for discussion, as well as the CNRS, the IRD, the ANR, and the Région Occitanie for funding (PHYEPI grant). This study was approved by the IRB of the CHU of Montpellier.

## Supplementary methods

### GLM

The main model was performed using a generalised linear model (GLM) assuming a binomial distribution, where the variable of interest was the binary variable *strain* (i.e. wild type or variant) and the explanatory variables were the sampling date (integer), the individual age (integer), the test kit used (boolean), the location of the sampling (boolean) and the region (factor). We further added an interaction between the region and the date.

Odds ratios were computed by estimating a likelihood profile.

We use a type II error for the analysis-of-variance given the uneven sampling between regions (the Anova function in the car package of R).

### Logistic growth fitting

We used the fitted values of the GLM model applied to the data after removing samples from hospitals (the sampling location effect was also obviously removed from the model) to perform the inference of a two-parameter logistic growth kinetic curve: 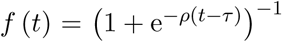, where *f* (*t*) is the frequency of the variants in the new infections at time *t, ρ* is the relative growth rate of the variants and *τ* is the time at which *f* reaches 1*/*2. This method is indeed more appropriate to deal with temporal auto-correlation biases in proportion time series [4, 5].

The parameter estimation was performed using the drc package in R both at the national and the regional level (for regions with at least 1,000 samples). The confidence intervals of the fitted curves rely on those of the estimated date of reaching half proportion of new infections (*τ*).

The unitless estimated transmission advantage, ETA, is expressed in terms of multiplicative gain in reproduction number with respect to that of the wild type, such that ℛ_variant_ = (1 + ETA) × ℛ_wild type_. Its calculation was made by solving the Euler-Lotka equation 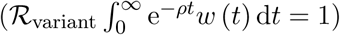 assuming a serial interval *w* following a Weibull distribution with a mean and SD of 4.8 and 2.3 days [10] and a constant ℛ_wild type_ equal to 1. The confidence interval rely on those of the estimated relative growth rate.

### Reproduction number and variant increase

The reproduction number was estimated using the method described in [11]. In brief, data was collected from https://www.data.gouv.fr/fr/datasets/donnees-hospitalieres-relatives-a-lepidemie-de-covid-19/ and the reproduction number was then computed for each French regions using the EpiEstim package in R by setting the serial interval to that reported by [10].

In order to estimate the increase in the proportion of variants among positive tests, we performed a GLM with a binomial distribution to explain the type of infection (wild type or variant) as a function or two factors (sampling date and individual age). This was done only on data collected outside hospital settings. The regression coefficients were used to perform a Spearman’s rank correlation with the most recent time point reproduction number estimate (that from Feb 14, 2021).

## Supplementary figures

**Figure S1:**
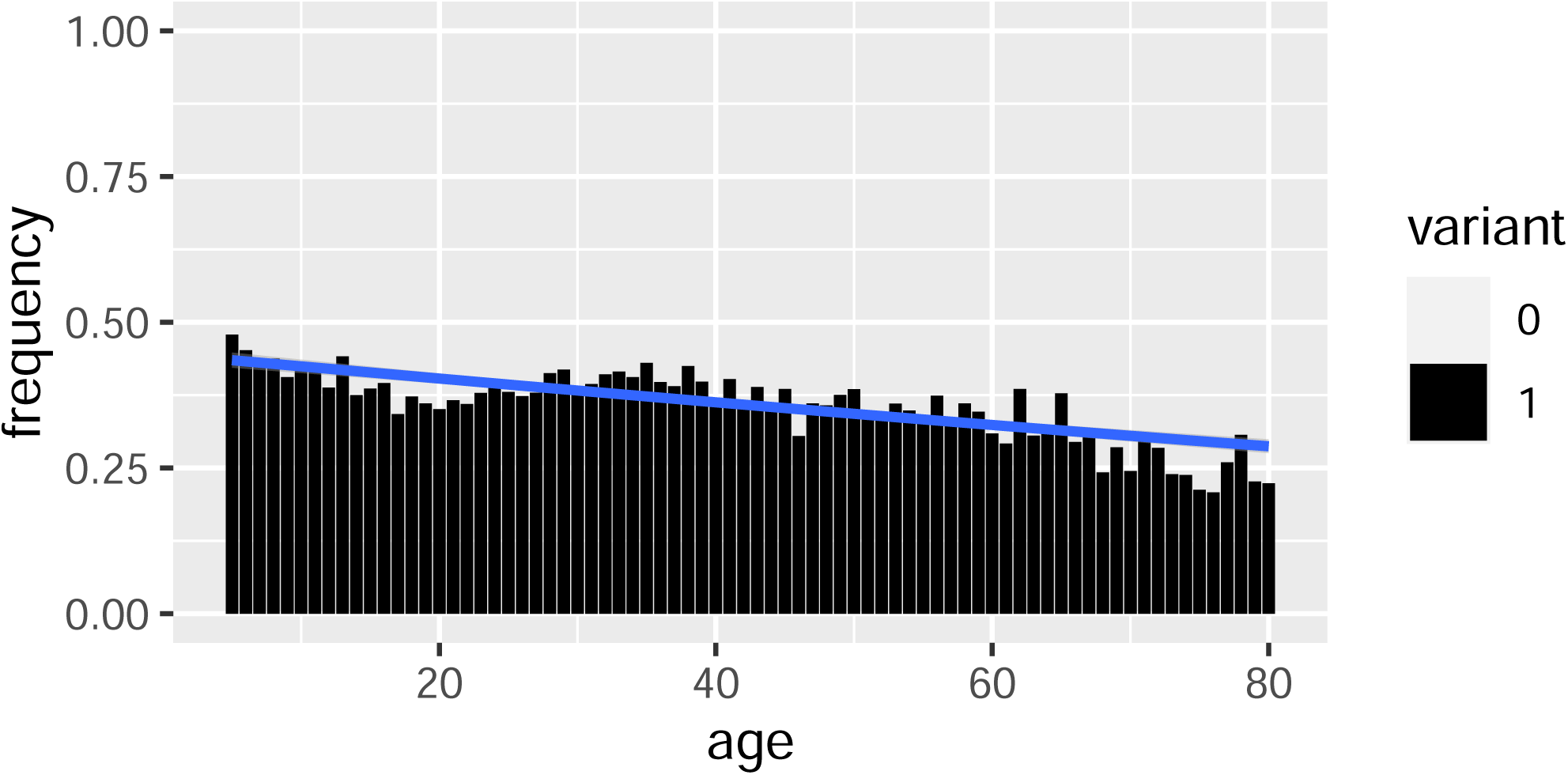
Proportion of infections caused by variants as a function of age. Uninterpretable tests are not shown.

**Figure S2:**
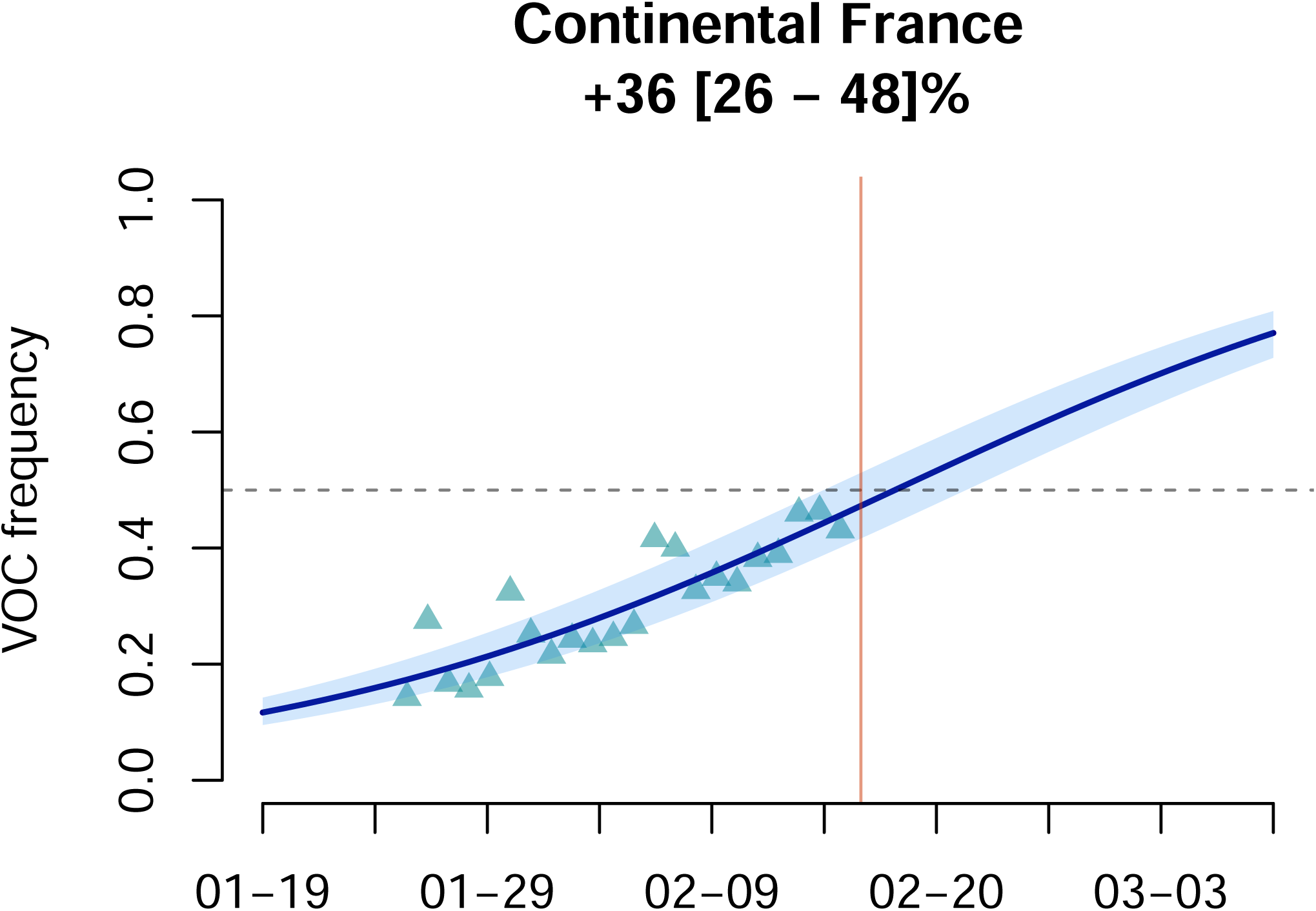
Variants frequency in France assuming that uninterpretable tests are all caused by ‘wild type’ strains.

**Figure S3:**
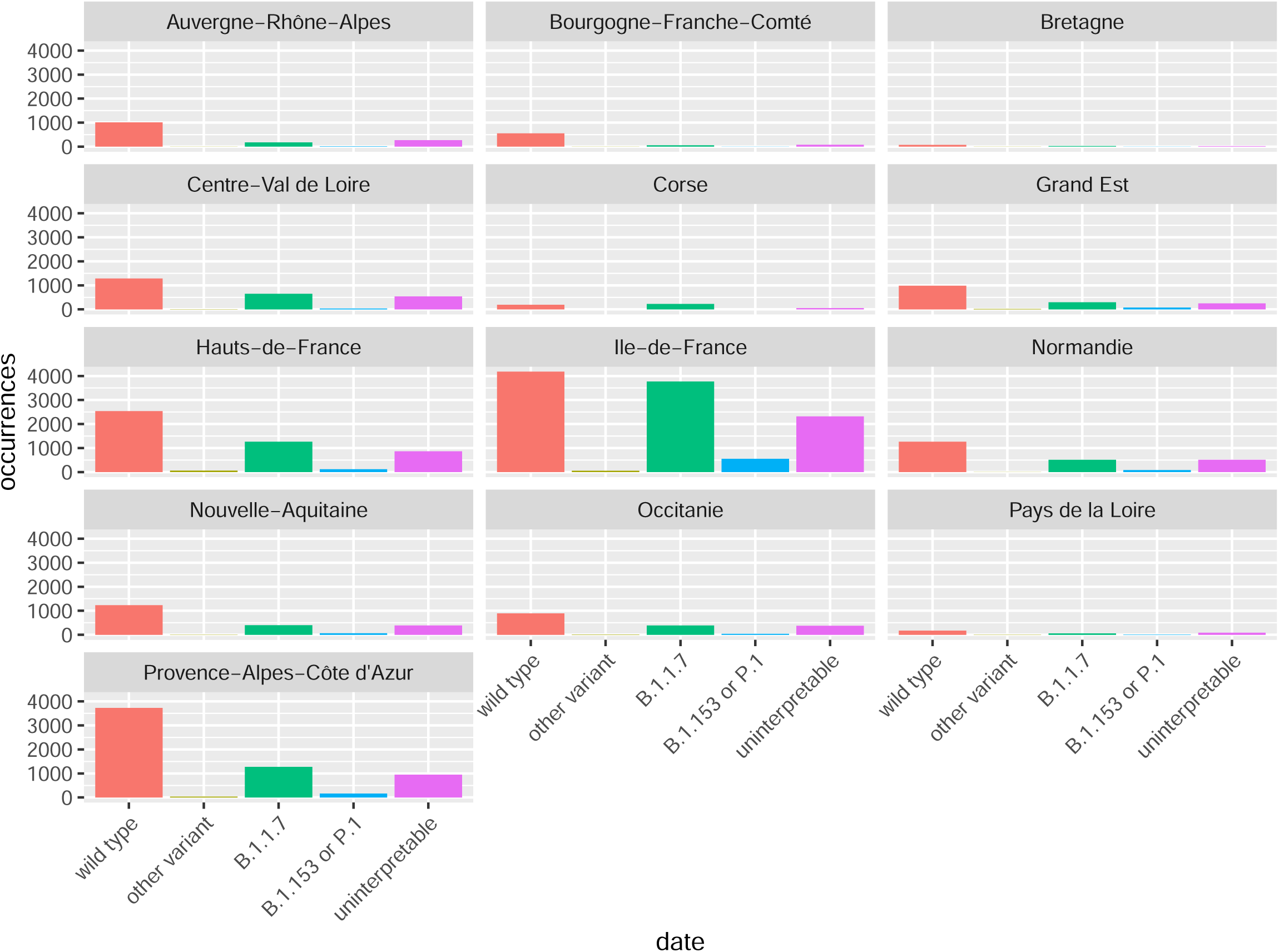
Outcome of variant-specific RT-PCR tests per region.

**Figure S4:**
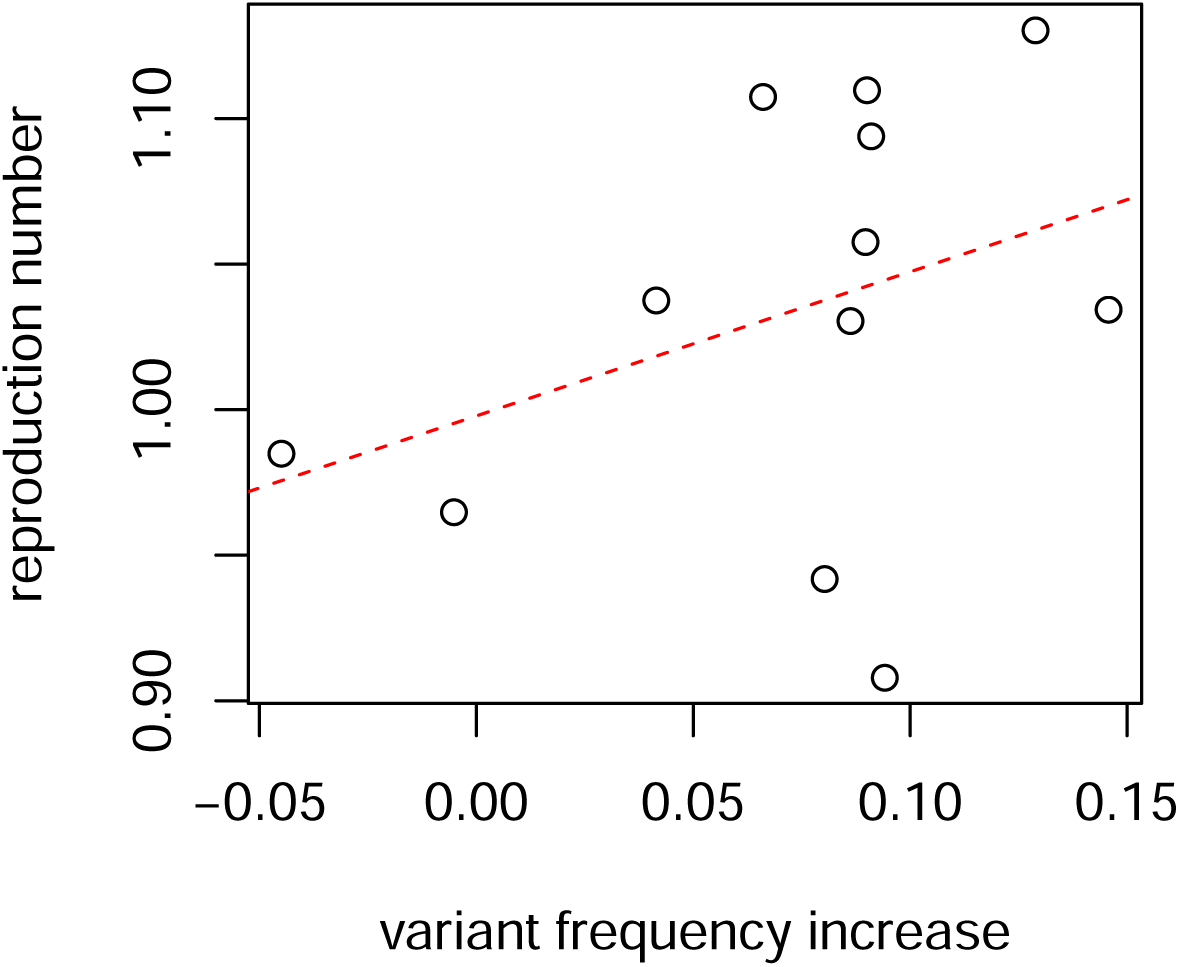
Regional reproduction number as a function of the estimated increase in variant frequency. The dashed line shows the output of a univariate generalised linear model.

## Notes

### Competing Interest Statement

The authors have declared no competing interest.

### Clinical Trial

NCT04738331

### Funding Statement

The authors than the CNRS, the IRD, the ANR and the Occitanie region (PHYEPI project) for funding.

### Author Declarations

This study has been approved by the IRB of the CHU of Montpellier (France).

